# Interpretable Machine Learning to Improve Donor-Recipient Matching at Time of Heart Transplantation

**DOI:** 10.64898/2026.07.29.26359283

**Authors:** Jie Xu, Wei Dai, Jason Goldberg, Inchi Hu, Chun-Hung Chen, Palak Shah, Christopher deFilippi, Jiayang Sun

**Affiliations:** George Mason University; INOVA Health System; University of Maryland, School of Medicine

**Keywords:** heart transplantation, data curation, interpretable machine learning, donor-recipient matching

## Abstract

**BACKGROUND:** Machine learning (ML) models have been used to evaluate one-year post-transplant mortality in donor-recipient pairs. Previous modeling utilizing non-interpretable ML methods (deep neural networks [DNN] and XGBoost) showed modest gains in area under the receiver operating curve (AUC) beyond logistic regression, but suffered a significant drop in predictive AUC applied to the subsequent years’ data and lacked statistical significance in validating previously identified risk factors.

**METHODS:** Using balanced SRTR datasets, we evaluated non-interpretable models against interpretable ML models (adaptive logistic regression with interaction terms [aLR], Classification and Regression tree [CART], and conditional inference tree [CIT]) for one-year mortality, including comprehensive clinician-supervised data curation and inclusion of variables describing pre- and post-2018 listing status changes. Models were trained/tested using rolling-window validation across years and further analyzed with repeated ten-fold cross-validation. Interaction terms were obtained via Adaptive Best-Subset Selection (ABESS).

**RESULTS:** Predictive validation before the listing policy change in 2018 showed similar AUCs between DNN (0.579), aLR (0.642), CART (0.579), and CIT (0.584), with XGBoost having a higher (0.763) AUC. However, in the post-2018 predictive analysis, aLR outperformed XGBoost (AUC 0.613 vs. 0.586). The interpretable ML models confirm the significance of previously reported risk factors (recipient bilirubin and creatinine) and identify risk factors not previously reported (donor pH, potential recipient distance, and recipient transfusion), as well as clinically relevant interaction terms.

**CONCLUSIONS:** Carefully developed interpretable ML models of one-year transplant mortality have similar predictive performance to black-box models, while identifying novel risk factors, and showing improved performance after recent listing policy changes. With appropriate validation and additional data, interpretable ML modeling may allow real-time data-driven donor selection.

## Introduction

Recent years have seen an increase in the number of U.S, adult heart transplants (HTx) from 1,974 in 2011 to 3,373 in 2021^1,2^. Over the same period, waitlist mortality has decreased from 15 deaths to 8.8 deaths per 100 person-years^2^, and one-year post-HTx mortality rates also decreased from 10.1% to 9.2%. However, the percentage of unused donor hearts remains high, with a current organ yield of ∼30%^2^. Various strategies have been studied to address this donor-recipient gap, including using machine learning (ML) to improve donors-recipient match ^3–5^. Yet, there remain major obstacles for ML models as HTx decision-support tools. Earlier studies using logistic regression^6–8^ showed limited predictive power for one-year post-Htx mortality, with Area Under the receiver operating characteristic Curve (AUC) values between 0.59 and 0.61 using United Network for Organ Sharing (UNOS) and International Society for Heart and Lung Transplantation (ISHLT) registry data^9,10^. Recent studies used “black-box” methods such as deep neural networks (DNNs), XGBoost, and Random Forest (RFs), which lack interpretability^1^. These models resulted in a modest increase in AUC beyond previous logistic regression methods, to approximately 0.65, but suffered a significant drop in AUC when applied to subsequent years’ data. In addition to their non-interpretable nature, these methodologies are prone to both overfitting and a lack of statistical significance in validation of specific risk factors^3,9–11^ beyond widely accepted predictors. Beyond predictive power, a practical ML model should be interpretable and built on well-curated real-world evidence to yield actionable insights for decision support. To address these issues, we built interpretable ML models using carefully curated data from the Scientific Registry of Transplant Recipients (SRTR) datasets, with the goal of identifying pre-transplant donor and recipient factors related to HTx outcomes.

## Methods

### Patient population and Outcome

With expedited IRB approval from George Mason University, we acquired SRTR standard analytics file (SAF) data augmented with donor-recipient match position data for HTx between January 2006 and March 2022. Our **primary analysis** focused on a cohort of 26,232 adults (>=18 years old, excluding those with multi-organ transplant) between January 2006 and October 2018 with 22,451 post-HTx survivals and 3781 deaths. This represents a *contemporary cohort* before the last major change on October 18, 2018 to the Organ Procurement and Transplant Network (OPTN) heart allocation policy from a three-tier system to a six-tier listing status system. We chose post-HTx **one-year mortality** as primary outcome. In our **second analysis**, we included 8,670 adult HTx between November 2018 and March 2021, with a one-year post-HTx follow-up record.

### Data curation

Among all 1,107 available variables, we removed variables irrelevant to outcome (e.g., IDs), unavailable at the time of donor-recipient matching (e.g., post-HTx information), or with near-zero variances. Missing values are addressed using our “divide-and-conquer-rejoin” (**DACr**) algorithm^12^. Briefly, categorical variables were regrouped or recoded as clinically sensible to address infrequent levels, and missing values were coded as their own level, “missing”. Numerical variables with >=10% missing values were *replaced* with surrogates with more complete information*, or excluded* if non-essential or redundant. *Imputation* was done for the remaining variables determined to be missing at random (MAR). The rest were recoded as categorical variables with an extra level of “missing”. The minimally missing subset (<10% per variable) actually had a maximum of <1% missing per variable, did not appear informative for the outcome, and accounted for <3% of total subjects. These subjects were thus removed. All steps were jointly reviewed by clinicians and statisticians, representing a data-driven, *domain-semi-supervised* filtering and missing-data treatment approach. We then used single-tree sure-screening and conditional tree to further reduce to 75 variables for final modeling building, which included donor and recipient variables, both widely used in previous analysis and variables not shown before, including recipient hemodynamics, histocompatibility crossmatch, and organ match run position.

Since the vast majority of HTx recipients survived the first year, this observational cohort is highly unbalanced between cases (deaths) and controls. Thus, we balanced controls with existing cases using propensity scores^12,13^. Furthermore, we exclude cases without the required ‘last status’ or blood type for donor–recipient matching, as the quality of these data may be questionable when the status is missing. All analyses were done using R 4.1.1.

### Model building and validation

Three interpretable ML models were constructed: 1) adaptive logistic regression (aLR) with meaningful interaction terms, 2) Classification and Regression tree (CART), and 3) conditional inference tree (CIT). Interaction terms were obtained via Adaptive Best-Subset Selection (ABESS)^14^. CART and CIT hyperparameters (complexity, split criteria, surrogate variables) were tuned on cross-validated predictive relative errors. The predictive performance was evaluated in two ways. First, we evaluated their performance using 01/2006-12/2016 training data and 01/2017-10/2018 test data, comparing these results to DNN and XGBoost models using the same datasets. This choice balanced the need to have sufficient data for training (10 years) and testing (2 years). Second, we conducted an expanding, rolling-window validation for the interpretable models: 1) train initial models using 2006 data and test models on 2007 data; 2) expand training dataset to include 2007 and test them on 2008 data; 3) continue adding subsequent year (up to 2017) data to training dataset and test each resulting trained model on the following year (up to 10/2018). This approach systematically validates interpretable models over time.

Our second analysis added a binary variable for the OPTN policy: three-tier (01/2006-10/2018) vs. six-tier (11/2018-03/2021) listing status. For aLR, the second analysis also experimented with adding interaction terms between this binary variable and other variables. Repeated ten-fold cross-validation (**RCV**) was used for model validation. Our model testing using available data before and after a significant policy change in 2018 has implications for model stability studies under future policy shifts, such as the October 2025 allocation changes.

## Results

### Patient population

Using propensity scores to match cases and controls on age and gender and excluding those without the required ‘last status’ and blood type for donor–recipient matching, our data curation approach produced a balanced (50-50% survival/deceased) sample of 5,732 HTxs with measures on 75 variables for the 01/2006-10/2018 cohort. The second cohort (11/2018-03/2021) had a balanced sample size of 1,151. We summarized donor and recipient variables identified as important by our interpretable models in **Table 1**, with column 2 reporting summary statistics for the raw sample and columns 3, 4, and 5 summarizing the balanced samples from 01/2006-12/2016, 01/2017-10/2018, and 11/2018-03/2022.

**Table 1:** Summary statistics of donor and recipient variables identified as important risk factors in the study cohort, balanced cohorts in 2006-2016, 2017-2018, and in 2019-2021.

| Key Variables<br>(abbreviation, unit) | Raw Data (n=26,232) | 2006-2016 (n=4624) | 2017-2018 (n=1108) | 2019-2021 (n=1151) |
| --- | --- | --- | --- | --- |
|  | median (1<sup>st</sup>, 3<sup>rd</sup><br>quartile) | median (1<sup>st</sup>, 3<sup>rd</sup><br>quartile) | median (1<sup>st</sup>, 3<sup>rd</sup><br>quartile) | median (1<sup>st</sup>, 3<sup>rd</sup><br>quartile) |
| Donor Age (dAge) | 31.67 (23.92, 41.92) | 31.08 (22.67, 43.08) | 31.46 (23.98, 41.17) | 31.92 (24.42, 40.83) |
| Recipient Age (rAge) | 56.50 (46.92, 63.25) | 56.58 (46.67, 63.17) | 57.75 (48.31, 64.25) | 59.92 (50.66, 65.17) |
| Recipient Bilirubin(rBili) | 0.7 (0.5, 1.1) | 0.8 (0.5, 1.2) | 0.7 (0.5, 1.0) | 0.7 (0.4, 1.1) |
| Recipient Creatine (rCreat) | 1.20 (0.96, 1.50) | 1.21 (1.00, 1.60) | 1.23 (1.00, 1.57) | 1.21 (0.97, 1.52) |
| Recipient Total IschemicTime(rHrIsch) in minutes | 190 (145, 229) | 196 (151, 236) | 191 (144, 232) | 206 (172, 242) |
| Recipient BMI (rBMI) | 27.0 (23.8, 30.7) | 27.02 (23.73, 30.74) | 27.51 (23.99, 31.40) | 27.74 (24.11, 31.45) |
| Donor PH (dPH) | 7.42 (7.38, 7.46) | 7.42 (7.37, 7.46) | 7.42 (7.38, 7.46) | 7.42 (7.38, 7.46) |
| Donor Ejection Fraction (dEjFract) | 60 (55, 65) | 60 (55, 65) | 60 (57, 65) | 60 (57, 65) |
| Recipient Time on Waiting List (rWaitTime, days) | 91 (27, 257) | 89 (25, 258) | 104 (30, 314) | 31 (8, 176) |
| abs(Predicted Heart Mass<sup>13</sup> -1) | 0.11 (0.05, 0.19) | 0.11 (0.05, 0.19) | 0.11 (0.05, 0.18) | 0.11 (0.05, 0.19) |
| PTR Distance (PTRDist, nautical miles) | 87.1 (12.3, 279) | 90.00 (13.55, 281.58) | 88.94 (14.04, 292.73) | 223.40 (84.45, 393.60) |
| Candidate Diagnosis: | **frequency (%tage)** | **frequency (%tage)** | **frequency (%tage)** | **Frequency (%tage)** |
| Ischemic | 9217 (35.1%) | 1761 (38.08%) | 327 (29.51%) | 360 (31.28%) |
| Nonischemic | 13295 (50.7%) | 2169 (46.91%) | 591 (53.34%) | 585 (50.83%) |
| Other | 3720 (14.2%) | 694 (15.01%) | 190 (17.15%) | 206 (17.90%) |
| Donor Troponin I level |  |  |  |  |
| Normal (< 0.4) | 13401 (50.53%) | 2265 (48.98%) | 707 (56.95%) | 636 (55.26%) |
| Abnormal (>= 0.4) | 4485 (17.26%) | 786 (17.00%) | 214 (17.15%) | 196 (17.03%) |
| Missing | 8346 (32.21%) | 1573 (34.02%) | 324 (25.90%) | 319 (27.72%) |
| Recipient Infection req. IV Drug Therapy 2 Weeks Prior to Tx |  |  |  |  |
| No | 23539 (89.7%) | 3978 (86.03%) | 993 (89.62%) | 1016 (88.27%) |
| Yes | 2693 (10.3%) | 646 (13.97%) | 115 (10.38%) | 135 (11.73%) |
| Recipient Ventilator Support |  |  |  |  |
| No | 20912 (79.72%) | 3565 (77.10%) | 889 (80.23%) | 978 (84.97%) |
| Yes | 5320 (20.28%) | 1059 (22.90%) | 219 (19.77%) | 173 (15.03%) |
| Recipient Had Transfusion |  |  |  |  |
| No | 18890 (72.01%) | 3261 (70.52%) | 832 (75.09%) | 982 (85.32%) |
| Yes | 7342 (27.99%) | 1363 (29.48%) | 276 (24.91%) | 169 (14.68%) |
| Recipient VAD Type |  |  |  |  |
| None | 14922 (56.88%) | 2653 (57.37%) | 545 (49.19%) | 711 (61.77%) |
| LVAD | 9800 (37.36%) | 1572 (34.00%) | 516 (46.57%) | 393 (34.14%) |
| Unspecified | 1510 (5.76%) | 399 (8.63%) | 47 (4.24%) | 47 (4.08%) |

### Predictive performance

**Figure 1** plots the ROC curves for the 01/2017-10/2018 test dataset, using aLR, CART, CIT, DNN, and XGBoost trained 01/2006-12/2016 data. **Table 2** shows training and testing AUCs achieved by these five methods: trained on 01/2006-12/2016 data and tested *before* (01/2017-10/2018) and *after* (11/2018-03/2021) the policy change. **Table 2** also shows AUCs of second analysis using combined data (01/2006-03/2021) and including a prior/post 2018 policy change binary indicator as well as an additional aLR model trained with interactions between the binary indicator and other variables. **XGBoost** and **DNN** had the top two **fitted** AUC values (0.828 and 0.707 respectively), with aLR trailing as the 3^rd^ (0.655) followed by CIT (0.627), and CART(0.648). **aLR and XGBoost** had the top-performing **testing** AUCs with aLR outperforming XGBoost (0.613 vs 0.586) on the post-policy-change data, and achieving similar results when adding the binary pre-post policy variable and the interactions. DNN’s AUC values in the primary analysis had a significant drop from 0.707 for training to (0.579, 0.581) for testing pre/post policy change. XGBoost’s AUC values dropped from 0.828 to (0.763, 0.586) and from 0.686 to 0.645, respectively, which is lower than DNN but higher than aLR. This analysis suggests that XGBoost and DNN are more sensitive to temporal shifts than the interpretable models.

**Figure 1:**
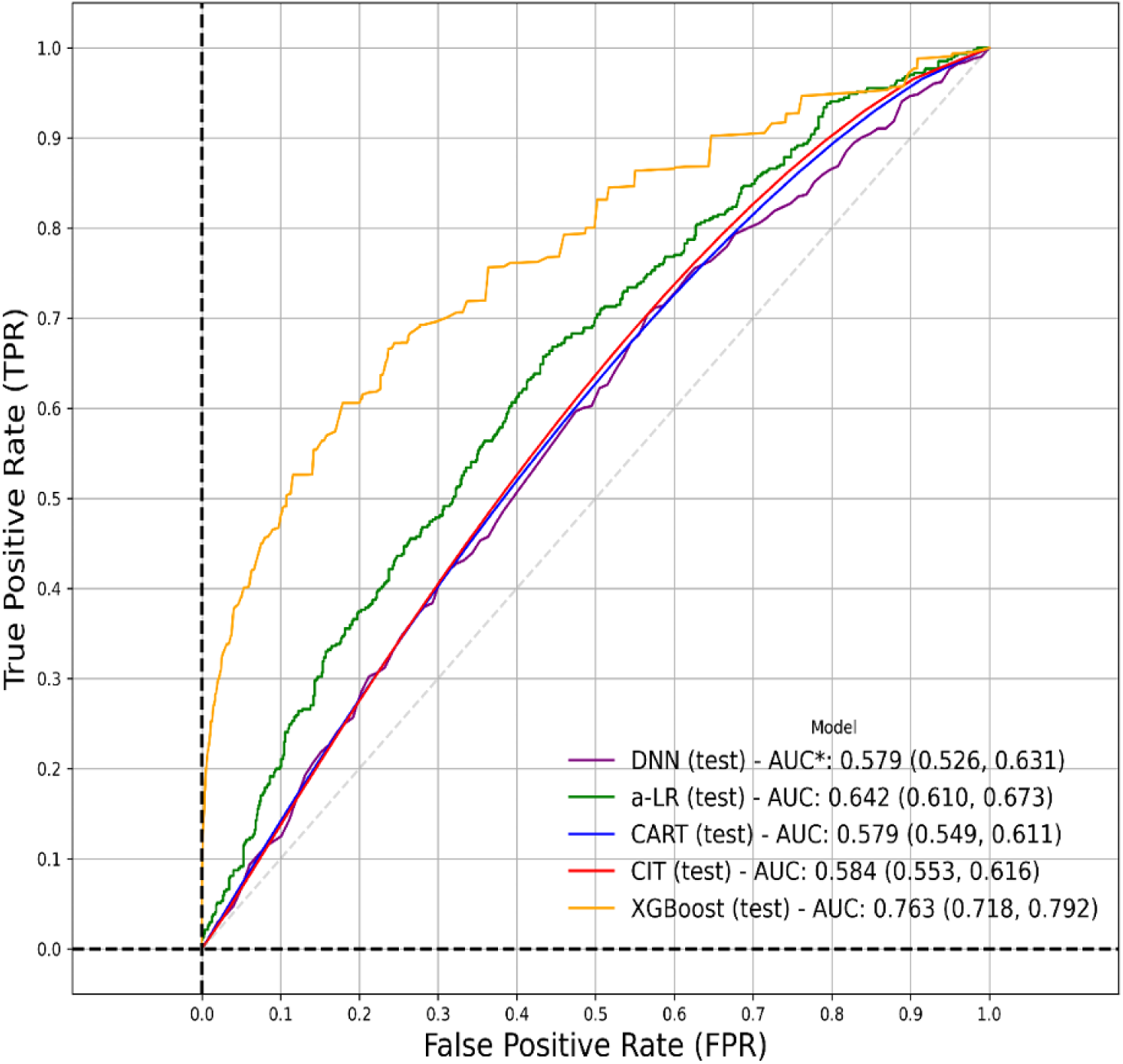
ROC curves with AUCs and 95% CI of AUC of CART, CIT, aLR, and DNN (NN) models trained on 2006-2016 data and tested on 2017-2018 data.

**Table 2:** AUCs achieved by trained ML models on different training and test datasets. **A1**: models trained using 2006-2016 data. **A2-1:** models trained using 01/2006-03/2021 data with a binary indicator to indicate if the HTx date was prior to or after 10/2018. **A2-2:** an additional aLR model trained with interactions between the binary indicator and other variables.

| Model – Data Performance Metric | A1: Fitted/ Training AUC: (2006-2016) | A1: Test AUC: 01/2017-10/2018 | A1: Test AUC: 11/2018-03/2021 | A2-1: Fitted/Training AUC | A2-2 Repeated CV (test) AUC |
| --- | --- | --- | --- | --- | --- |
| aLR | 0.655 | 0.642 | 0.613 | 0.648/0.654 | 0.643/0.641 |
| CART | 0.648 | 0.579 | 0.559 | 0.623 | 0.571 |
| CIT | 0.627 | 0.584 | 0.571 | 0.632 | 0.597 |
| DNN | 0.707 | 0.579 | 0.581 | 0.726 | 0.590 |
| XGBoost | 0.828 | 0.763 | 0.586 | 0.686 | 0.645 |

### Rolling-window performance

**Figure 2** presents AUC values of the interpretable models from the rolling window validation. Starting from 2008, test data AUC values were close to training AUC values, with the exception of 2014-2016 test AUC. The differences are relatively minimal, with the largest drop observed for aLR in year 2013 (AUC 0.65) to 2014 (AUC 0.57). We did not perform rolling-window validation for DNN and XGBoost models because they require a much larger sample size than the interpretable models to establish the first reliable training model to start rolling validation. However, Table 2 shows the significant AUC drops of XGBoost and DNN on test data from subsequent years. As an **external** validation, the best AUC reported in Miller et al.^3^ dropped from 0.893 (with RF) to 0.657 (with XGBoost) when model validation was changed from cross-validation to rolling-window validation.

**Figure 2:**
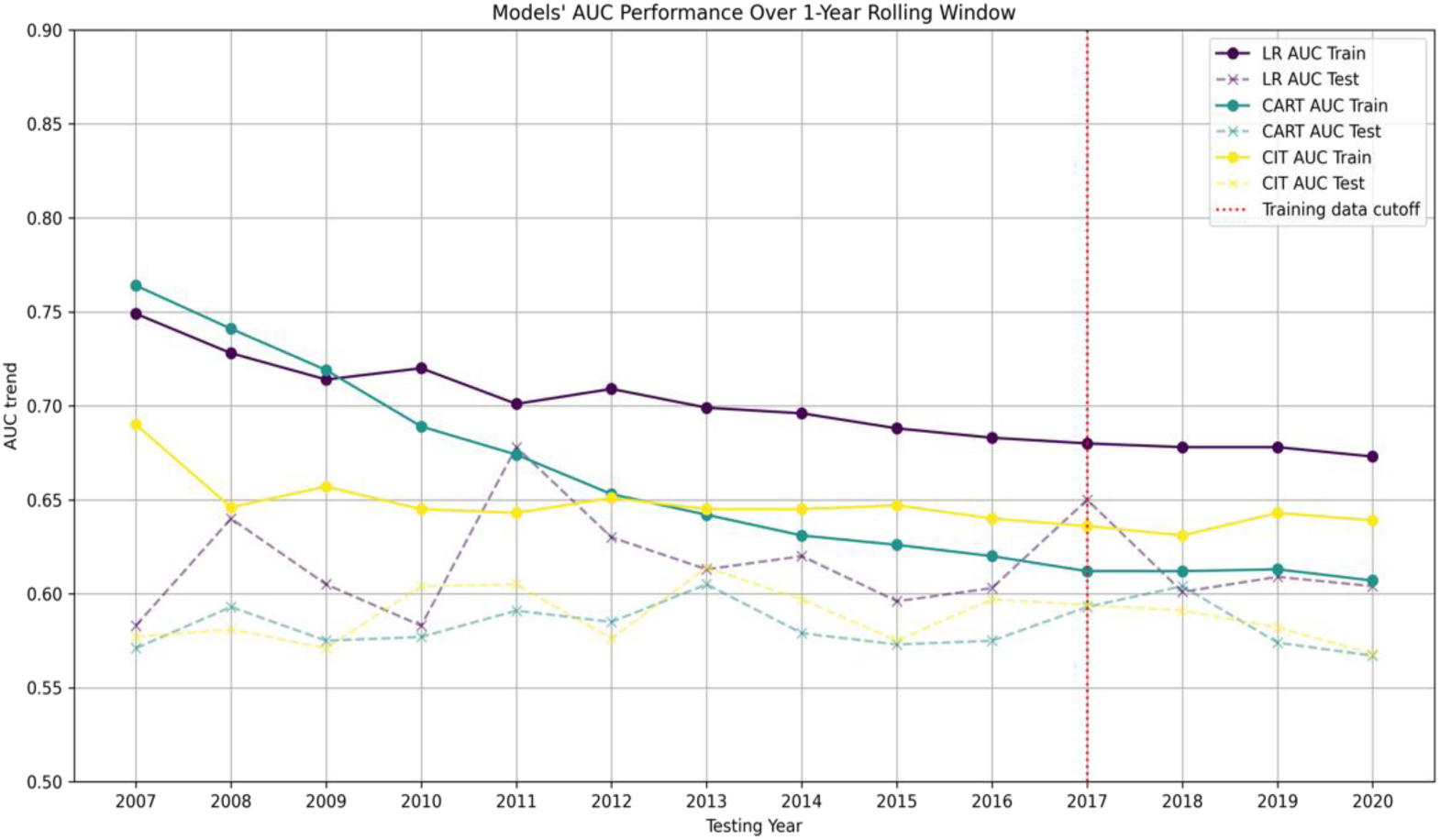
ROC curves of CART, CIT, and aLR models using rolling validation. The year on the horizontal axis indicates the last year covered by the training data. The vertical axis gives the AUC value for the test data in the following year. For example, the AUC value mapped to the year 2008 was for the model trained on 2006-2008 data and tested on 2009 data.

### Interpretable ML models and risk factors identified

**Table 3** reports the aLR model trained from the primary analysis on 01/2006-10/2018 data. This model confirms the significance of widely cited risk factors, e.g., recipient bilirubin and creatinine^3,6,7,10,11^. We also identified significant risk factors not previously reported in the literature: donor pH, potential transplant recipient (PTR) distance, and recipient had transfusion prior to HTx. Furthermore, we identified four significant interaction terms including: 1) recipient used hypertension drug & ventilator support, 2) recipient had a transfusion prior to HTx & received treatment for infection prior to surgery, 3) recipient had a diagnosis of non-ischemic cardiomyopathy & donor-recipient race was White-White, and 4) recipient BMI & recipient had a diagnosis other than ischemic or non-ischemic cardiomyopathy. **An interaction term is considered significant when both conditions linked by “&” co-occur, forming a subgroup with a significantly different risk than other groups.**

**Table 3:**
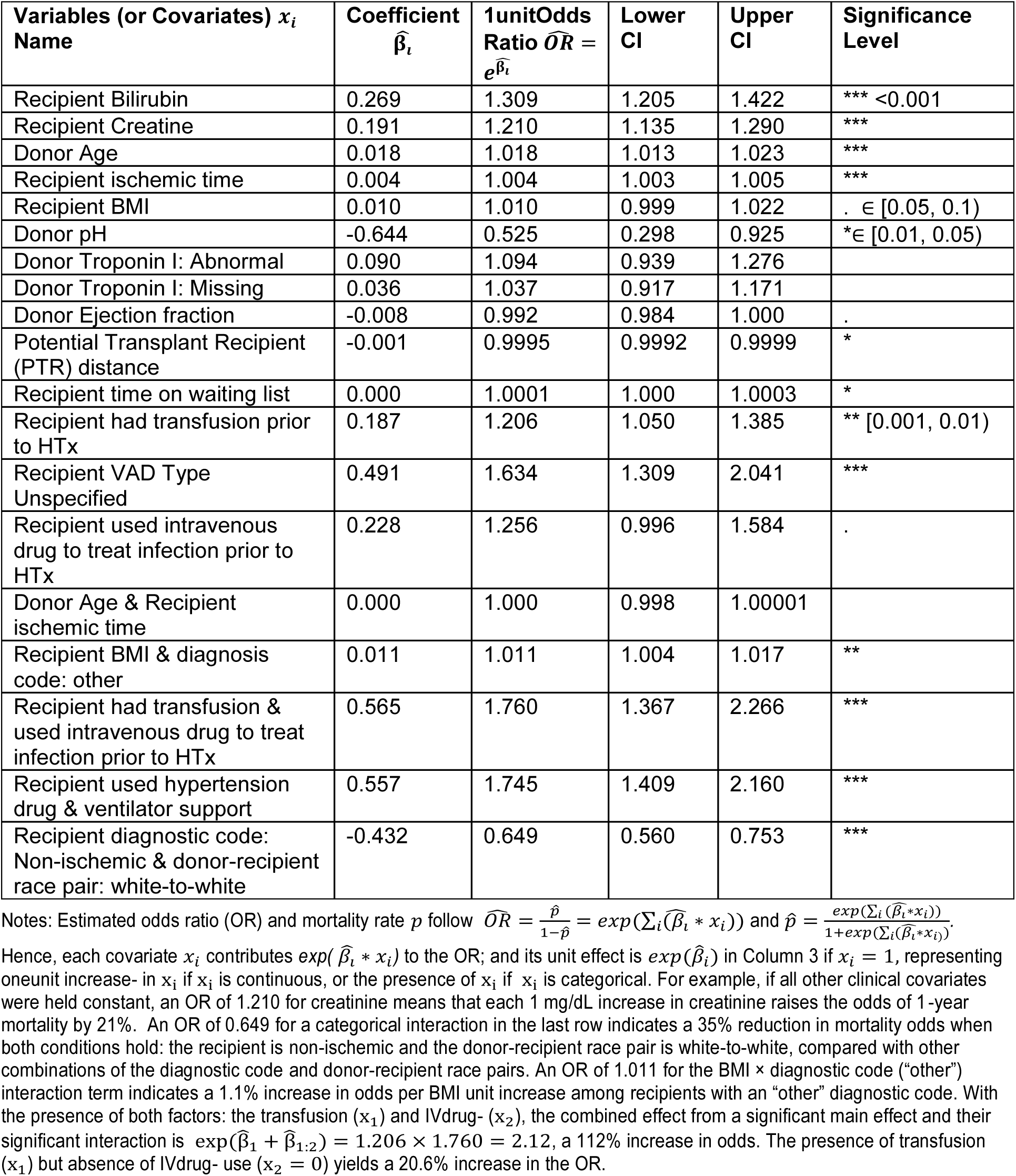
Important variables for predicting one-year post-transplant mortality identified by adaptive logistic regression aLR. Interaction terms are indicated by “&”. The blue italic fonts indicate variables not reported as significant in previous literature. Signif. Codes: 0 “***” 0.001 “**” 0.01 “*” 0.05 “.” 0.1 “ “

| Variables (or Covariates) $x_i$<br>Name | Coefficient<br>$\hat{\beta}_i$ | 1unitOdds<br>Ratio $\widehat{OR} = e^{\hat{\beta}_i}$ | Lower<br>CI | Upper<br>CI | Significance<br>Level |
| --- | --- | --- | --- | --- | --- |
| Recipient Bilirubin | 0.269 | 1.309 | 1.205 | 1.422 | *** <0.001 |
| Recipient Creatine | 0.191 | 1.210 | 1.135 | 1.290 | *** |
| Donor Age | 0.018 | 1.018 | 1.013 | 1.023 | *** |
| Recipient ischemic time | 0.004 | 1.004 | 1.003 | 1.005 | *** |
| Recipient BMI | 0.010 | 1.010 | 0.999 | 1.022 | . $\in [0.05, 0.1)$ |
| Donor pH | -0.644 | 0.525 | 0.298 | 0.925 | * $\in [0.01, 0.05)$ |
| Donor Troponin I: Abnormal | 0.090 | 1.094 | 0.939 | 1.276 |  |
| Donor Troponin I: Missing | 0.036 | 1.037 | 0.917 | 1.171 |  |
| Donor Ejection fraction | -0.008 | 0.992 | 0.984 | 1.000 | . |
| Potential Transplant Recipient (PTR) distance | -0.001 | 0.9995 | 0.9992 | 0.9999 | * |
| Recipient time on waiting list | 0.000 | 1.0001 | 1.000 | 1.0003 | * |
| Recipient had transfusion prior to HTx | 0.187 | 1.206 | 1.050 | 1.385 | ** $[0.001, 0.01)$ |
| Recipient VAD Type Unspecified | 0.491 | 1.634 | 1.309 | 2.041 | *** |
| Recipient used intravenous drug to treat infection prior to HTx | 0.228 | 1.256 | 0.996 | 1.584 | . |
| Donor Age & Recipient ischemic time | 0.000 | 1.000 | 0.998 | 1.00001 |  |
| Recipient BMI & diagnosis code: other | 0.011 | 1.011 | 1.004 | 1.017 | ** |
| Recipient had transfusion & used intravenous drug to treat infection prior to HTx | 0.565 | 1.760 | 1.367 | 2.266 | *** |
| Recipient used hypertension drug & ventilator support | 0.557 | 1.745 | 1.409 | 2.160 | *** |
| Recipient diagnostic code: Non-ischemic & donor-recipient race pair: white-to-white | -0.432 | 0.649 | 0.560 | 0.753 | *** |

Notes: Estimated odds ratio (OR) and mortality rate $p$ follow $\widehat{OR} = \frac{\hat{p}}{1-\hat{p}} = \exp(\sum_i (\hat{\beta}_i * x_i))$ and $\hat{p} = \frac{\exp(\sum_i (\hat{\beta}_i * x_i))}{1 + \exp(\sum_i (\hat{\beta}_i * x_i))}$ .
Hence, each covariate $x_i$ contributes $\exp(\hat{\beta}_i * x_i)$ to the OR; and its unit effect is $\exp(\hat{\beta}_i)$ in Column 3 if $x_i = 1$ , representing oneunit increase- in $x_i$ if $x_i$ is continuous, or the presence of $x_i$ if $x_i$ is categorical. For example, if all other clinical covariates were held constant, an OR of 1.210 for creatinine means that each 1 mg/dL increase in creatinine raises the odds of 1-year mortality by 21%. An OR of 0.649 for a categorical interaction in the last row indicates a 35% reduction in mortality odds when both conditions hold: the recipient is non-ischemic and the donor-recipient race pair is white-to-white, compared with other combinations of the diagnostic code and donor-recipient race pairs. An OR of 1.011 for the BMI × diagnostic code (“other”) interaction term indicates a 1.1% increase in odds per BMI unit increase among recipients with an “other” diagnostic code. With the presence of both factors: the transfusion ( $x_1$ ) and IVdrug- ( $x_2$ ), the combined effect from a significant main effect and their significant interaction is $\exp(\hat{\beta}_1 + \hat{\beta}_{1:2}) = 1.206 \times 1.760 = 2.12$ , a 112% increase in odds. The presence of transfusion ( $x_1$ ) but absence of IVdrug- use ( $x_2 = 0$ ) yields a 20.6% increase in the OR.

**Figures 3 and 4** plot the CART and CIT models trained from the primary analysis. Both models confirm the significance of previously reported variables (recipient bilirubin, recipient creatinine, total ischemic time, donor age)^3,6–11^. They also show a new risk factor (recipient had a transfusion prior to HTx) for certain subgroups of patients. For CART, recipient creatinine >=1.615 was the strongest discriminator of 1-year post-HTx mortality. For CIT, recipient bilirubin > 2 is the strongest discriminator of 1-year post-Htx mortality. We compared the variables identified by aLR, CART, and CIT with a Venn diagram in **Figure 5**, with the following variables shared by all three models: recipient bilirubin, total ischemic time, recipient transfusion status, recipient creatine, and donor age.

**Figure 3:**
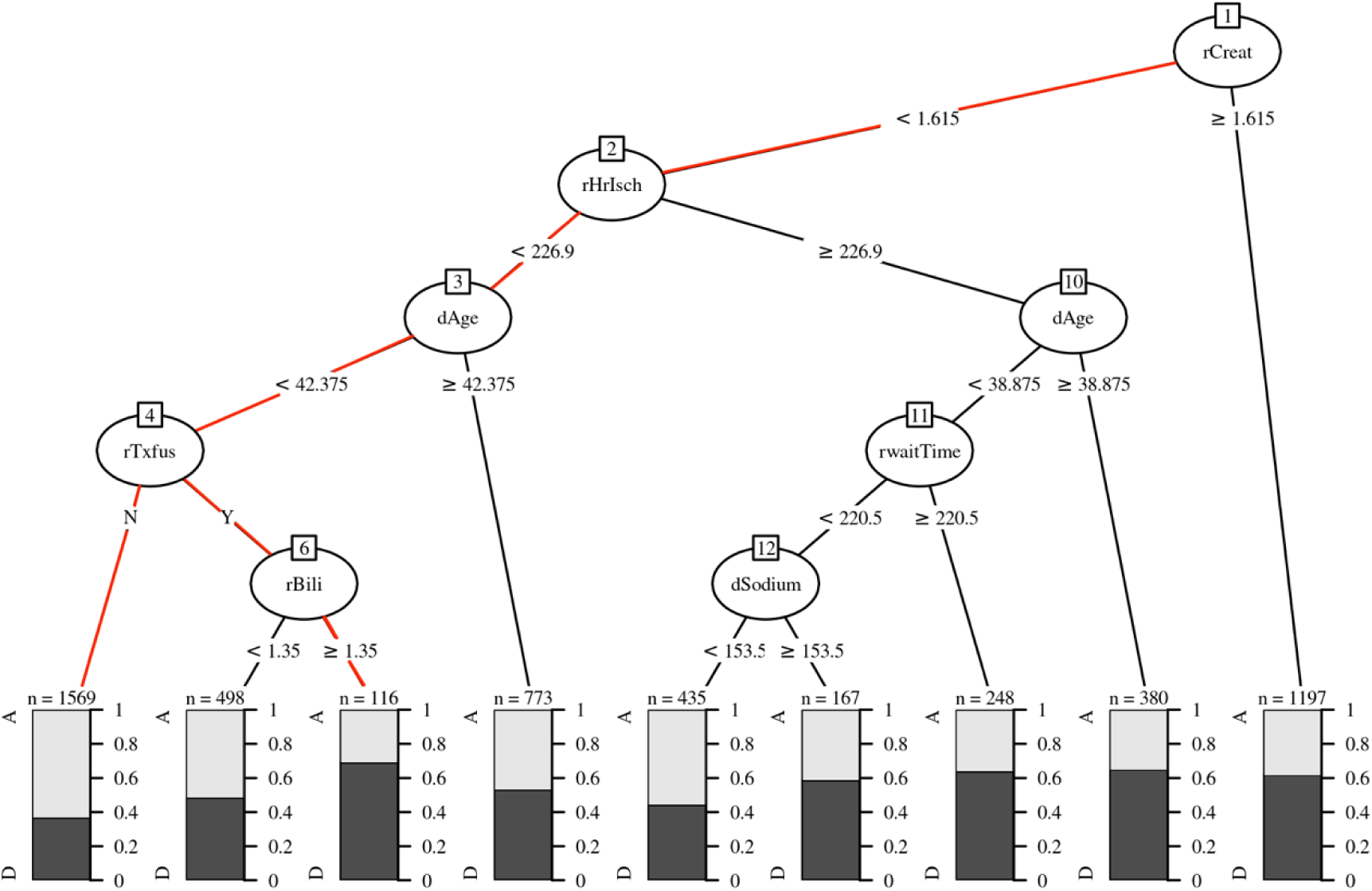
Classification tree (CART) to predict one-year post-HTx mortality. The red branch shows the conditions that lead to the two most predictive patient populations. For each leaf node, the dark bar indicates the percentage of “Death” outcomes for the patient population in this node, with the sample size shown above the bar. **rCreat:** recipient’s most recent serum creatinine (mg/DL); **rHrisch:** total ischemic time (minutes); **rTxfus:** recipient transfusion before HTx; **rBill:** recipient total bilirubin (mg/dL); **rwaitTime**: time from recipient added to the waiting list to HTx (days); **dAge**: donor age (years), **dSodium**: last serum sodium prior to organ procurement (mEq/L).

**Figure 4:**
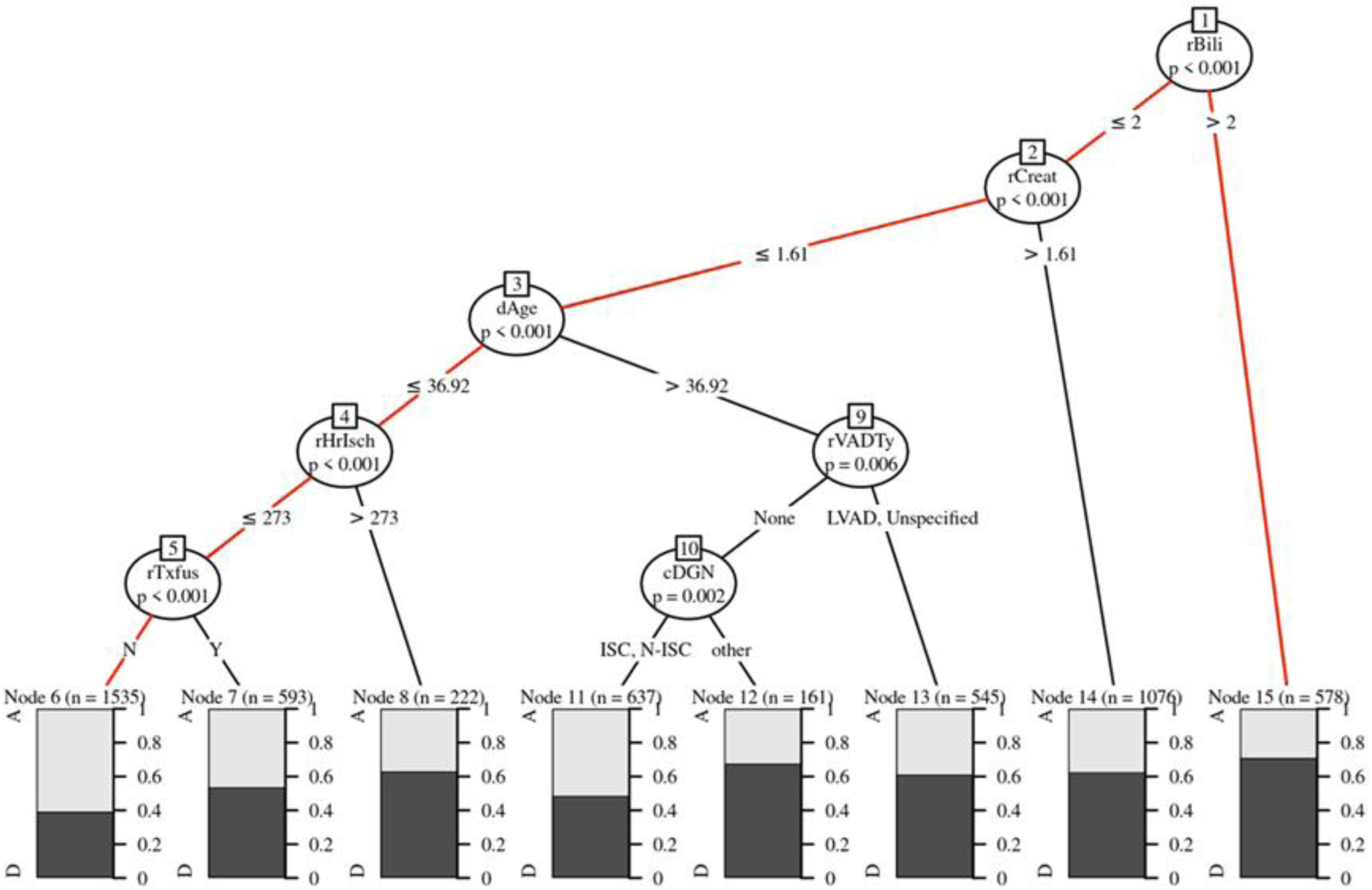
Conditional inference tree (CIT) to predict one-year post-HTx mortality. The red branch shows the conditions that lead to the most predictive patient populations. For each leaf node, the dark bar indicates the percentage of the “Death” outcomes for the patients population grouped into this node, with the sample size given above the bar. **rBill**: recipient total bilirubin (mg/dL); **rCreat**: recipient most recent serum creatinine (mg/DL); **dAge**: donor age (years); rHrisch: total ischemic time (minutes); **rTxfus**: recipient transfusion before HTx; **rVADTy**: type of ventricular assist device patient was on; **cDGN**: patient primary diagnosis (ISC: ischemic, N-ISC: nonischemic, other: other diagnosis such as congenital).

**Figure 5:**
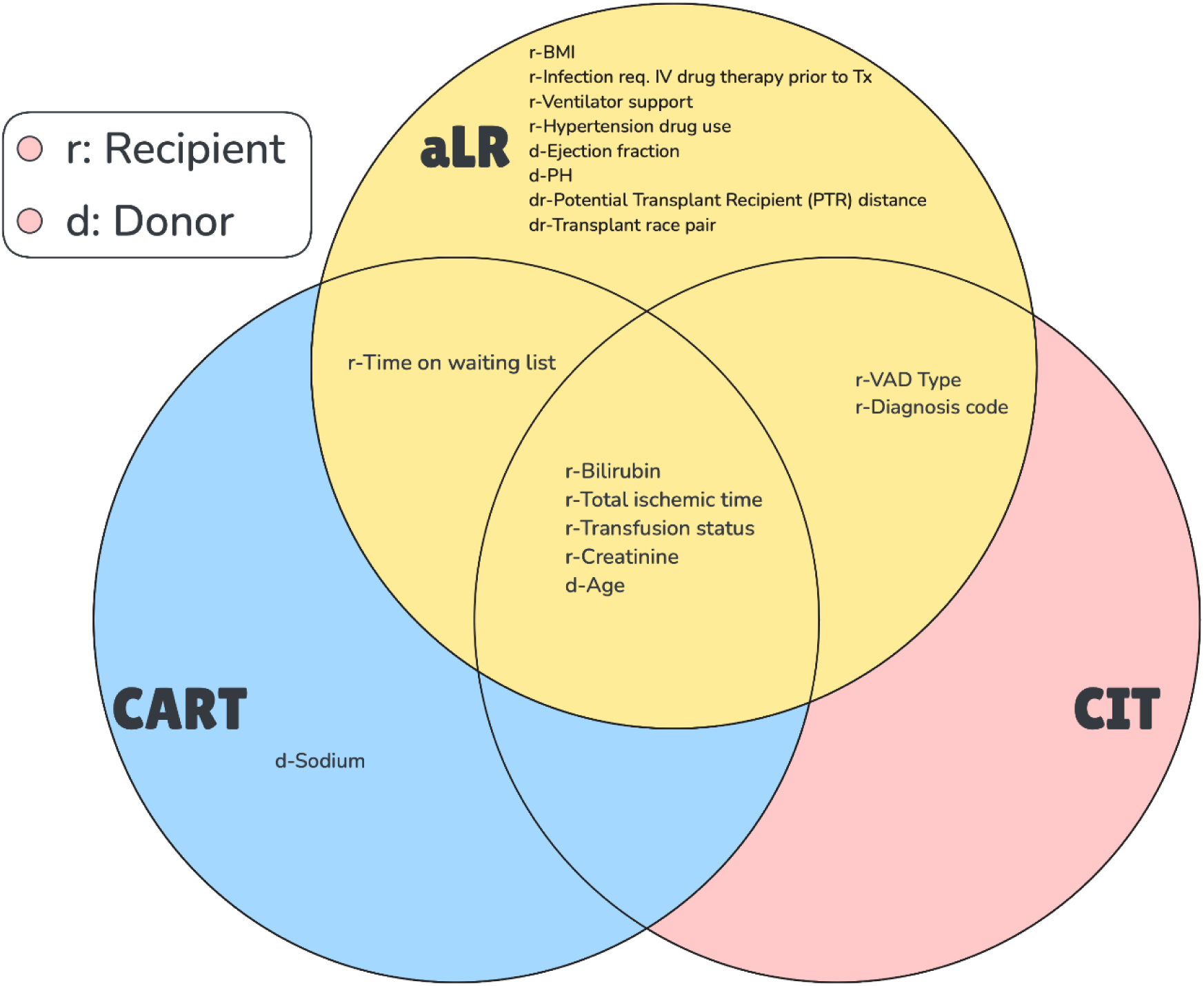
Venn Diagram of Donor and recipient variables identified as important risk factors for one-year post-HTx mortality by logistic regression (LR), CART, and CIT.

Our second analysis shows that the effect of the binary policy year indicator is significant, indicating a higher one-year mortality risk in 11/2018-03/2021, with an odds ratio of 1.222 at a significance level of 0.05 in the aLR model. When interaction terms between the policy year indicator and other variables are considered, the only significant factor involving the policy year indicator is its interaction with recipient BMI, with an odds ratio of 1.039 at a significance level of 0.05. The CIT model shows higher risk in 2018-2021 for a subgroup of patients with no transfusion prior to surgery, lower ischemic time (< 227 minutes), younger donor (<=42.83 years old), lower recipient creatinine (<1.79), and nonischemic cardiomyopathy, **Table 1**, there was a slight increase in recipient BMI over the years. Overall, the increased post-2018 risk may be linked to higher BMI, amplified during the COVID-19 pandemic, and potentially confounded by its effects in some subgroups.

In terms of model stability, **RCV** was used to validate the performance of different models. Our testing of the models’ performance using available data before and after a significant policy change in 2018 has implications for model stability studies under future policy shifts^2^.

## Discussion

This study derived three interpretable models that are competitive with DNN/XGBoost models, based on SRTR data, by developing a systematic, domain-semi-supervised data-driven process for data curation. Our models not only confirmed the significance of widely reported risk factors but also identified new clinically meaningful variables and interaction terms and provided variable thresholds. The models derived herein can inform explainable point-of-care tools to evaluate a donor-recipient potential match.

### Previous donor-recipient one-year post-HTx Mortality Risk Models

We compared our work with previous studies that predicted one-year post-transplant mortality in supplement Table 1. Compared to previous studies, our study cohort was strategically chosen to contain a contemporary cohort under the same allocation policy. Our study confirmed the importance of donor and recipient age, recipient total bilirubin, recipient creatinine, ischemic time, and recipient BMI that were reported by multiple previous studies as important risk factors^3,6–11^. Our models did not explicitly report recipient age as an important risk factor, as it was a significant risk factor in our primary analysis and thus used as a factor in propensity score-based matching. We identified several new variables not reported previously: recipient blood transfusion prior to HTx, donor pH, PTR distance, and donor ejection fraction. While donor ejection fraction is clinically known as an important variable in clinician’s decisions, it has not been identified as a statistically significant variable in past studies, likely due to the lack of variability in ejection fraction values in the data. Our aLR showed that ejection fraction was a useful factor in predicting survival, but with a small effect size and marginal significance, confirming the above finding. Furthermore, our aLR model identified 4 statistically significant interaction terms, which have important clinical relevance. For example, recipients both a) receiving hypertension medications and requiring ventilator support or b) being treated for infection and having blood transfusion, representing subgroups with more significant mortality risk than those with each factor alone. Other interaction terms raise question for future study, such as the risk among recipients with non-ischemic cardiomyopathy having a same race transplant. These newly discovered factors and interaction terms describe drivers of post-transplant mortality that may allow for better risk prediction in donor-recipient matching.

### Novel Modeling and Validation

Because tree-based models such as CART and CIT make minimal assumptions, they are useful for validating results from parametric models and for offering additional insights beyond linear models. In addition, tree models provide thresholds of these risk factors, specifically for recipient bilirubin and creatinine, which may aid in clinical decision making. The CART and CIT models also provide insights on how additional risk factors may be incorporated to enable HTx deemed too risky otherwise. For example, **Figure 3** shows that a donor-recipient match including recipient creatinine < 1.615, waitlist time <220.5 days, and a young donor (<39 years) with adequate serum sodium level (<153.5 mEq/L) may mitigate the risk of long ischemic time (>226.9 minutes), which has mortality risk of ∼0.4 in this scenario, much lower than others at 0.6 or greater.

Figure 2 shows the AUCs on the rolling-window validation data were close to the AUCs on the training data, attesting to the generalization performance of interpretable models over time and the potential for the stability of these models in future patient populations. For the CART and CIT models, there were larger differences in training and test AUCs than those of aLR due to less training data early on. For 2015 and 2016 test data, the aLR model showed a bigger drop in AUCs. To investigate if this drop is due to a temporal shift, we conducted a correlation analysis between the model coefficients and the AUCs across the validation cohorts, which showed significant correlation among multiple variables’ coefficients, showing the presence of a temporal shift. There were several notable trends in this period, including s shifts in donor cause of death and the widespread adoption of LVAD.

### Clinical Implications

Currently, much of the practice of donor-recipient matching includes manual clinician application of previously identified risk factors, without the inclusion of evidence-based tools to evaluate post-transplant risk. As ML models continue to become available to clinicians, the results of this study may be applied to donor-recipient pairs and provide risk prediction, allowing the utilization of organs that previous methodology may not have deemed appropriate. Previous “black box” ML models have lacked clinical utilization, in part due to clinicians’ inability to determine which risk factors they included. Due to the interpretability of the identified models as well as their rigorous training and validation, these results can aid in the development of real-time decision support for clinicians. Our second analysis showed an increased risk of mortality after HTx in 11/2018-03/2021. This change may reflect a shift in the recipient population, likely partially related to changes in matching policy, and may be confounded by the COVID-19 pandemic. With additional data following the new policy change in Oct 2025, further insights may be gained.

## Conclusions

Our data curation and modeling pipeline suggest that interpretable ML models can achieve predictive performance comparable to the best performance reported in literature using ML models that do not offer the interpretability desired by clinicians. Our study provides insights on previously unknown risk factors and potential mitigation strategies against known risk factors. It also shows that careful modeling, such as including an indicator for policy changes, can help reduce the impact of data shifts from different periods.

The limitations of our study include the lack of detailed individual-level clinical data and genetic biomarkers, and data after the October 2025 policy change. Future access to and integration of these additional data may enable a more comprehensive model with higher AUCs and prove more applicable in the clinical setting.

## Data Availability

he data that support the findings of this study were obtained from the Scientific Registry of Transplant Recipients (SRTR) under a data use agreement. Restrictions apply to the availability of these data, and they are therefore not publicly available. Researchers may apply directly to SRTR for access to the underlying data. Analytic methods and supplementary materials are provided in the manuscript and online supplement.

https://srtr.hrsa.gov/requesting-data/about-srtr-standard-analysis-files-safs/

## List of Non-standard Abbreviations

UNOS: United Network for Organ Sharing
SRTR: Scientific Registry of Transplant Recipients
OPTN: Organ Procurement and Transplantation Network
AUC: area under the curve
ML: machine learning
ANN: artificial neural network
DNN: deep neural network
ISHLT: International Society of Heart and Lung Transplant
BMI: body mass index

## Acknowledgements

This research is supported in part by NSF-SCH Award IIS-2123683, INOVA Health U19-11-3826, and INOVA Health OSRPSD-4133.

## Supplementary materials

An online supplement provides detailed information on data sets, data curation, model training and validation, and additional analyses.

## Author contributions

Sun conceived the idea based on a different project led by Chen and co-led the project with Xu. Goldberg helped derive methodology, oversaw data curation, and contributed to manuscript preparations. Clinicians DeFilippi and Shah aided in data curation and manuscript editing. Statisticians Sun, Hu, and Dai, and Operations Researchers Chen and Xu collaborated on the entire research. Xu and Dai acquired the SRTR data under clinicians’ direction. Sun and Xu supervised data curation and analyses performed by Dai, who wrote algorithms and code. Xu drafted the 1^st^ version of the manuscript with heavy input from Sun. All authors reviewed, edited or provided feedback, and approved the manuscript.

## Footnotes

1 We distinguish interpretable and explainable models as follows. Interpretable models are transparent by design and can be directly understood without additional tools (e.g., linear and logistic regression, GLM, decision trees, and other physics-based or nonlinear parametric models). In contrast, explainable models rely on post hoc methods, such as SHAP values or LIME (Local Interpretable Model-agnostic Explanations), to provide feature attribution scores or local approximations of the behavior of otherwise complex and opaque models, such as these black-box models.

2 Including cases with missing ‘last status’ as a separate “missing” category increased the balanced sample to 7,562 cases for the 1/2006–10/2018 period. Modeling results using 5,732 versus 7,562 cases were nearly identical, which is expected, as ‘last status’ was not an important predictor in any model in Table 2 with complete information. The aLR model with 7,562 cases showed a slightly lower AUC, likely reflecting poorer data quality in cases missing the last status.

