## Supplementary Tables and Figures for "Interpretable Machine Learning to Improve Donor-Recipient Matching at Time of Heart Transplantation"

**SUPPLEMENTAL Figure A**

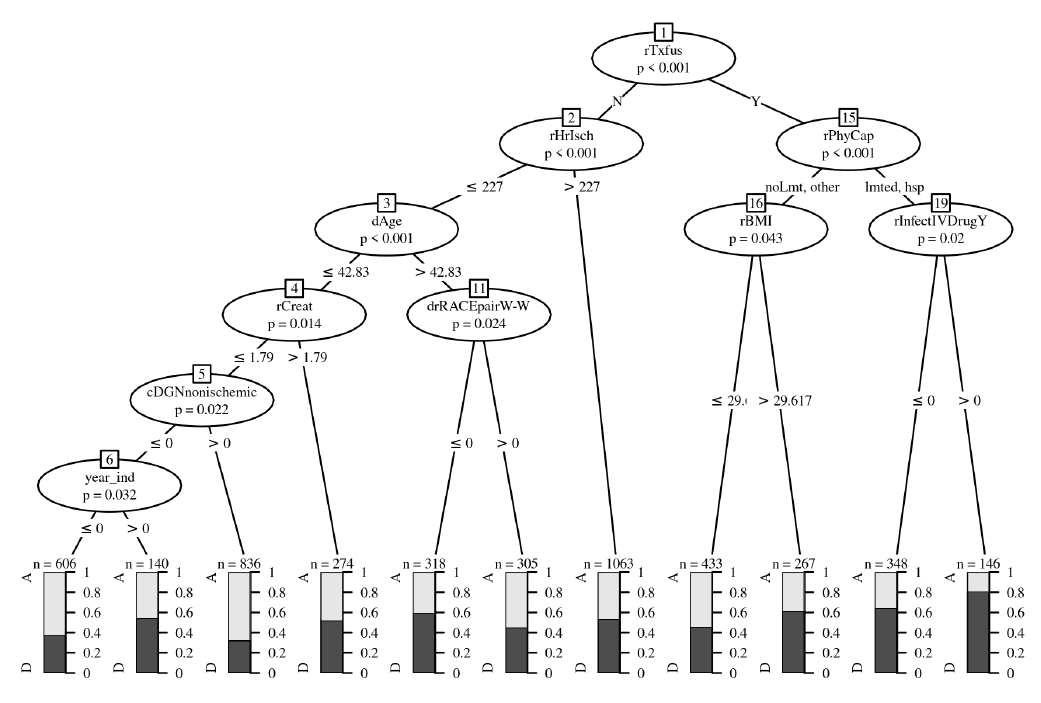

**Figure A:** Conditional inference tree trained on 01/2006-03/2021 data with binary policy year indicator.

**SUPPLEMENTAL TABLES**

**Supplemental Table 1.** Comparison of this work and previous works on predicting adult post-transplant one-year mortality

| **Paper** | **Study Cohort** | **Models** | **Reported Model Performance (AUC)** | **Important Variables (when multiple models were used, only variables identified by at least two models are reported)** |
| --- | --- | --- | --- | --- |
| Hong et al. (2011)^7^ | UNOS data, Jan 2001 - Dec 2007, 11,703 adult HTx recipients | Multivariate logistic regression | - | Mechanical circulation support, renal failure, total bilirubin, donor and recipient age |
| Weiss et al. (2012)^8^ | UNOS data, 1996-2007, 22,252 adult HTx recipients | Multivariate logistic regression on donor variables | - | Ischemic time, donor age, race mismatch, donor BUN/creatinine ratio |
| Weiss et al. (2011)^6^ | UNOS data, 1997-2008, 21,378 adult HTx recipients | Multivariate logistic regression on recipient variables to create a recipient index (IMPACT) | 0.569 on 2010-2018 UNOS data as validated in Medved et al. (2018)^9^ | Recipient age, recipient bilirubin, recipient creatinine, heart failure etiology, recipient race, VAD |
| Nilsson et al. (2015)^11^ | ISHLT registry data, 1994-2010, 56,625 HTx | Artificial Neural Network (ANN) | 0.65 (derivation)  0.62 (validation), | Donor and recipient age, recipient creatinine, diagnosis, time era, infections within two weeks prior to HTx, gender, mechanical ventilation |
| Medved et al. (2018)^9^ | UNOS data, Jan 1997 - Dec 2011, 27,860 HTx | Comparison of IMPACT and IHTSA | IMPACT: 0.61 (test), 0.61 (derivation); IHTSA: 0.65 (test), 0.69 (derivation) | Gender, heart failure etiology, infection, mechanical ventilation (shared between IMPACT and IHTSA) |
| Kampaktsis et al. (2021)^10^ | UNOS data, 2010-2018, 18,625 recipients | AdaBoost, SVM, decision tree, kNN, logistic regression | AdaBoost: 0.689 (validation) | Recipient creatinine, recipient height, ischemic time, recipient BMI, year of transplant, type of VAD, recipient prior cardiac surgery, heart failure etiology, donor age, recipient age |
| Miller et al. (2022)^3^ | UNOS data, Jan 1994 - Dec 2016, 59,590 adult HTx recipients | Random Forest (RF), XGBoost, L2 regularized logistic regression | 10-fold CV: 0.893 (RF); Rolling CV: 0.657 (XGBoost) | Recipient bilirubin and creatinine (identified by all three ML models); any ventilation support, retransplant indication, donor heart biopsy performed (identified by logistic regression and random forest); congenital heart disease, total artificial heart, donor history of cancer, cause of death (identified by logistic regression and XGBoost), combined panel reactive antibody (PRA), recipient age, recipient creatinine at listing, recipient height, donor tattoos, donor vessels with stenosis, donor age, expanded criteria cardiac transplant, height ratio, ischemic time (identified by random forest and XGBoost). |
| This paper | SRTR data  Jan 2006-  Oct 2018  26232 adult HTx recipients | XGBoost,  Sure Independence Screening (SIS),  Decision trees,  logistic regression | logistic regression:  0.652 (testing data on 2017-2018) | Recipient bilirubin, recipient blood transfusion prior to HTx, recipient creatinine, recipient BMI, donor age (identified by logistic regression, CART, CIT), recipient age (identified by logistic regression and CART), recipient physical capacity (identified by CART and CIT) |

**Supplemental Table 2.** List of top 20 variables selected by Deep Neural Networks trained on January 2006 to December 2016 cohort (DNN-A1) according to permutation importance on January 2017 to October 2018 validation set with importance mean and standard deviation.

| Variable | Importance Mean | Importance Std |
| --- | --- | --- |
| Recipient Creatinine | 0.133959 | 0.006669 |
| PTR Distance (nautical miles) | 0.107437 | 0.005494 |
| Recipient Time on Waiting List (days) | 0.102860 | 0.004467 |
| Recipient Total Ischemic Time(minutes) | 0.096270 | 0.003260 |
| Candidate Primary Diagnosis: nonischemic | 0.069039 | 0.003153 |
| Recipient BMI | 0.065973 | 0.004073 |
| Partial pressure of oxygen (pO2) in donor lungs on a fraction of inspired oxygen (FiO2) | 0.057643 | 0.001967 |
| Recipient Pulmonary Artery Pressure Mean | 0.050389 | 0.002102 |
| Recipient Pulmonary Capillary Wedge (PCW) Pressure Mean | 0.031922 | 0.002169 |
| Recipient Bilirubin | 0.025240 | 0.002245 |
| Donor Age | 0.024577 | 0.003451 |
| Donor BUN | 0.018513 | 0.003019 |
| Donor pCO2 | 0.017002 | 0.002637 |
| Donor-Recipient Race Pair: White-to-White | 0.013753 | 0.002317 |
| Donor SGPT | 0.011442 | 0.002605 |
| Donor Ejection Fraction | 0.011236 | 0.001829 |
| Donor Cause of Death: Stroke | 0.010503 | 0.002174 |
| Recipient Used Intravenous Drug to Treat Infection prior to HTx: Yes | 0.010343 | 0.001627 |
| Donor Serum Sodium Level | 0.009451 | 0.002238 |
| Donor Blood Partial Pressure of Oxygen | 0.008810 | 0.001964 |

**Supplemental Table 3.** List of top 20 variables selected by XGBoost trained on January 2006 to December 2016 cohort (XGBoost-A1) according to permutation importance on January 2017 to October 2018 validation set with normalized importance scores.

| Variable | Normalized Importance Score |
| --- | --- |
| Recipient Creatinine | 100.00 |
| Recipient Total Ischemic Time(minutes) | 97.607195 |
| Donor Age | 88.145397 |
| Recipient Bilirubin | 53.776351 |
| Predicted Heart Mass Ratio | 41.765962 |
| Recipient Had Transfusion prior to HTx: Yes | 40.299871 |
| Recipient BMI | 34.009528 |
| Recipient Time on Waiting List (days) | 32.970733 |
| Recipient VAD Type Unspecified: Yes | 28.304058 |
| Donor Blood Partial Pressure of Oxygen | 26.515719 |
| PTR Distance (nautical miles) | 26.443331 |
| Recipient Used Intravenous Drug to Treat Infection prior to HTx: Yes | 26.159855 |
| Donor Blood PH | 25.727586 |
| Recipient Pulmonary Artery Pressure Mean | 24.651087 |
| Recipient Age | 24.201591 |
| Recipient Physical Capacity: Other | 24.190581 |
| Donor Ejection Fraction | 23.327309 |
| Donor-Recipient Race Pair: White-to-White | 23.217653 |
| Donor pCO2 | 22.655568 |
| Donor INR | 22.612205 |

**Supplemental Table 4.** Coding and frequency of recipient VAD type.

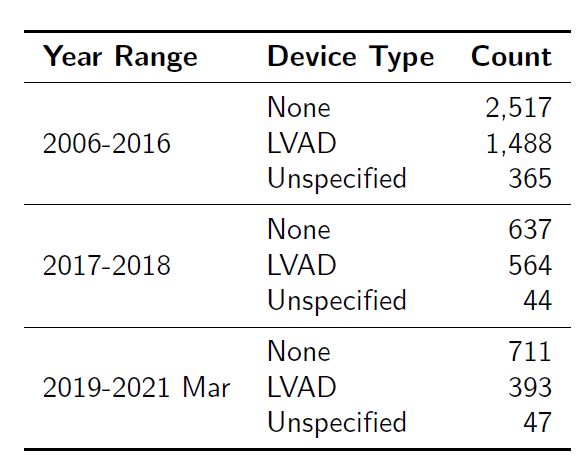

**Supplemental Table** 5. Candidate diagnosis code and frequency

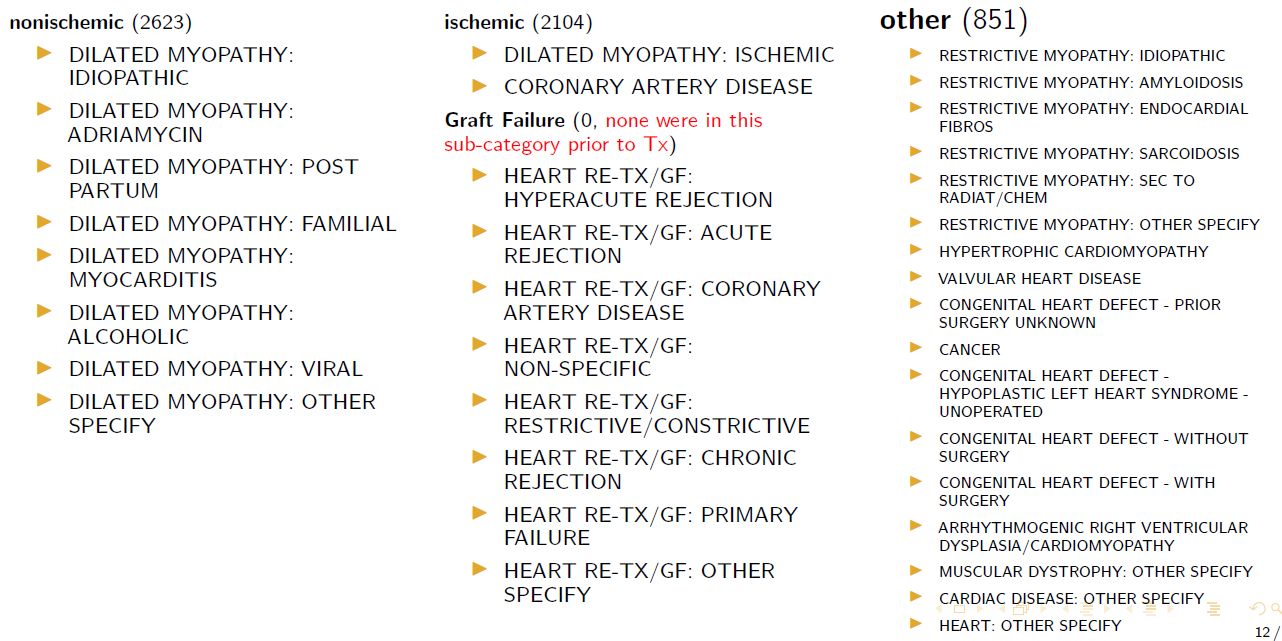

Supplemental Table **6.** Summary of significant interaction terms in aLR

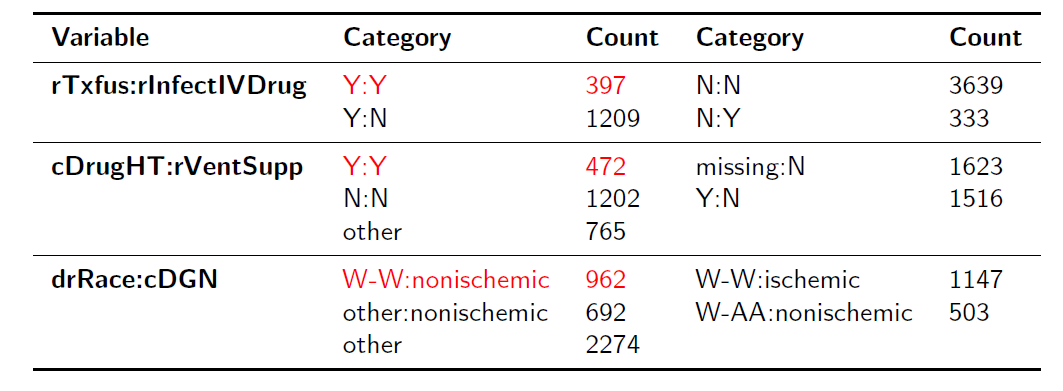
